# phenotypic and Genotypic Detection of Carbapenemases in Clinical Isolates of *Acinetobacter baumannii*

**DOI:** 10.1101/2024.11.20.24317647

**Authors:** Alya Amer Rahi

## Abstract

*Acinetobacter baumannii,* a Gram-negative coccobacillus with a short and round rod-like shape, is an opportunistic pathogen in immunocompromised patients, particularly prevalent as a nosocomial infection. A notable characteristic is its diverse antibiotic resistance mechanisms. The isolation of *A. baumannii* strains in hospitals is becoming more prevalent, presenting an escalating challenge in the treatment process. In the present study involving isolates obtained from diverse sources (blood, urine, wound, and burn swabs), the Antimicrobial Susceptibility Testing (AST) results for(28)*Acinetobacter baumannii* isolates revealed pronounced resistance. Notably, resistance rates were notably high for piperacillin (80%) and carbenicillin (81’.2%). Resistance to Imipenem and meropenem stood at 8.4% and 19.8%, respectively, while the highest resistance was recorded against gentamicin (82%), amikacin (72.6%), cefepime (60.8%), cefotaxime (70.2%), ceftazidime (70.2%), and ceftriaxone (71.6%). Molecular detection of Enzymatic genes was executed through PCR testing, revealing that, All 28 isolates (100%) exhibited the presence of the *bla_OXA-_*_51_type gene, a considered pointer for finding of bacteria by PCR. The prevailing MBL gene was *bla_VIM_*-type, identified in 12(42.8)% of isolates. Furthermore, 10(35.7%)of isolates carried the *bla_KPC_*_-_ gene. In conclusion the identification of these genes (*bla_OXA-51_*, *bla_VIM_,* and *bla_KPC_-*type) in *Acinetobacter baumannii*. Addressing antibiotic-resistant bacteria challenges healthcare; crucial to understand, monitor, and regulate antibiotic resistance gene dissemination for public health.

## 1 Introduction

*Acinetobacter baumannii* is a non-motile, Gram-negative bacterium that is well-known for causing infections acquired in hospitals. It demonstrates adaptability and resilience on a variety of surfaces, particularly in healthcare settings(1). Treatment becomes more difficult and multidrug-resistant strains of these bacteria can arise due to capacity to develop resistance to multiple antibiotics. People who have compromised immune systems are more susceptible to infections like wounds, bloodstream infections, and order to prevent nosocomial infections and manage public health issues, stringent infection control protocols are essential. (2). Strict infection control measures are essential to deal with public health issues and prevent outbreaks in hospitals. Rapid detection of Acinetobacter baumannii as an antibiotic-resistant strain is immediate, and a successful strategy should stop the spread of the bacteria by using the last antibiotic such as colistin to eliminate MDR-AB multidrug resistance – *A.baumannii*.(3).*Acinetobacter baumannii* has molecular enzymatic genes that are critical for immunity to antibiotics and the uptake of nutrients, especially iron, that are necessary for the bacterium to survive in its host. Due to their the enzymatic pathways of *Acinetobacter baumannii* is the focus of current research activities(4). By altering or breaking down antimicrobial agents, these enzymes present a serious problem in clinical settings.The way that the bacterium gets iron is closely related to the range of enzymes that it has. Treatment choices are restricted by the pathogenicity of iron, which is enhanced by enzymes that both scavenge and absorb iron. To successfully overcome these obstacles, creative therapeutic strategies are crucial(5). The focus of current research is on figuring out *Acinetobacter baumannii’s* enzymatic genes. This could ultimately lessen the impact of *Acinetobacter baumannii* in clinical settings by improving the management and treatment of infections caused by the bacteria.

## 2. Material and methodes

**1** **Clinical Specimens**:28 out of 400 (7%) isolates were collected from Hilla teaching hospital between February2023 to September2023.These specimens included blood (75), urine (75), wound swab samples (100),burn swab(150). The samples underwent culturing on MacConkey agar, chromogenic agar, blood agar, and brain heart infusion broth. Identification was carried out using PCR tests and the Vitek-2 system. The diagnostic process involved assessing characteristics, morphology, and microscopic examination with gram stain, adhering to CLSI-2023 guidelines. This comprehensive approach aimed to accurately identify and characterize pathogens present in diverse clinical specimens.
**2** Antimicrobial Susceptibility Testing (AST) using the Disk Diffusion Test (DDT) was conducted on 28(7%) *Acinetobacter baumannii* isolates. The testing involved a range of antibacterial agents, including Piperacillin, Ceftazidime, Cefotaxime, Carbenicillin, Ceftriaxone, Imipenem, Gentamicin, Amikacin, Ciprofloxacin, and Meropenem. The DDT (Disc Diffusion Test) procedure involved the cultivation of specimens on Muller-Hinton agar medium, followed by the addition of antibiotic discs. and incubation at **37°C** for reading the results Recorded outcomes were assessed by(CLSI,2023). This standardized methodology was the susceptibility of *A. baumannii* isolates to different antimicrobial agents.

### Ethical approval

The study was approved by the Ethics Committee of Al-Hillah Surgical Teaching Hospital, participants were fully informed about the objectives of the research, and their consent was obtained before collecting samples for experiments and publishing the results. Additional approval was granted by the local Ethics Committee of the College of Medicine, University of Babylon and the Hospital Ethics Committee under document no.[IRB: 6-20, 22/1/2023].

#### 1 DNA Extraction and PCR Test

DNA was extracted from bacteria using DNA extraction kits, and primers specific for gene detection in *A. baumannii* were employed based on the information provided in Table (1).

**Table (1).**
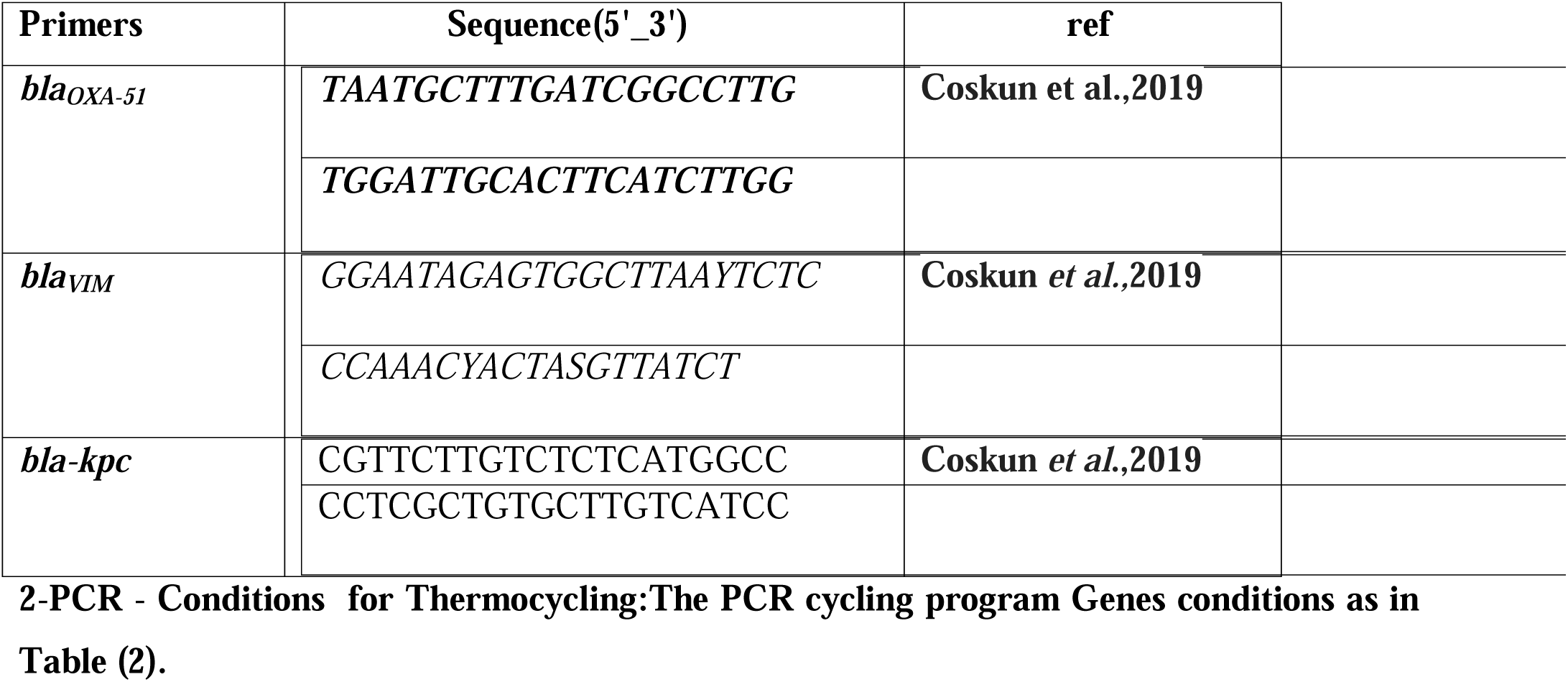
Primers used in this research.

**Table (2):**
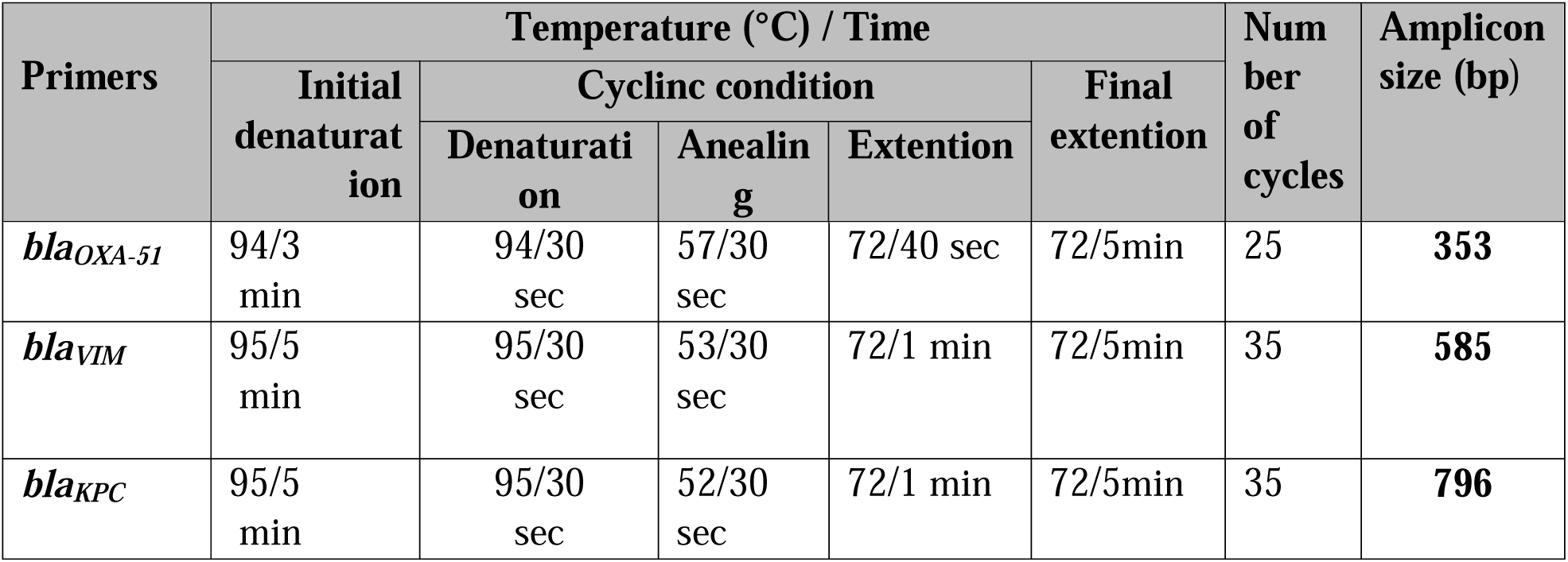
The conditions for genes detection used in this study.

## 3. Results and Discussion

A total of 28 clinical specimens of *Acinetobacter baumannii* were isolated from a pool of 400 specimens, encompassing blood (75), urine (75), wound swab samples (100), burn swab(150). This accounts for 7%% of the specimens, as represented in figure (1). All isolates were obtained from Hilla Teaching Hospital in Iraq, spanning the timeframe from February 2023 to September 2023. The isolation process involved culturing on various media, including MacConkey agar, chromogenic agar, blood agar, and brain heart infusion broth.

Identification criteria included the appearance of red colonies on chromogenic agar and positive results in biochemical tests (Simmons citrate agar +, catalase test positive). The diagnosis of all *Acinetobacter baumannii* isolates was confirmed using the VITEK2-system, ensuring a comprehensive and reliable assessment of these clinical specimens.

**Table (1):**
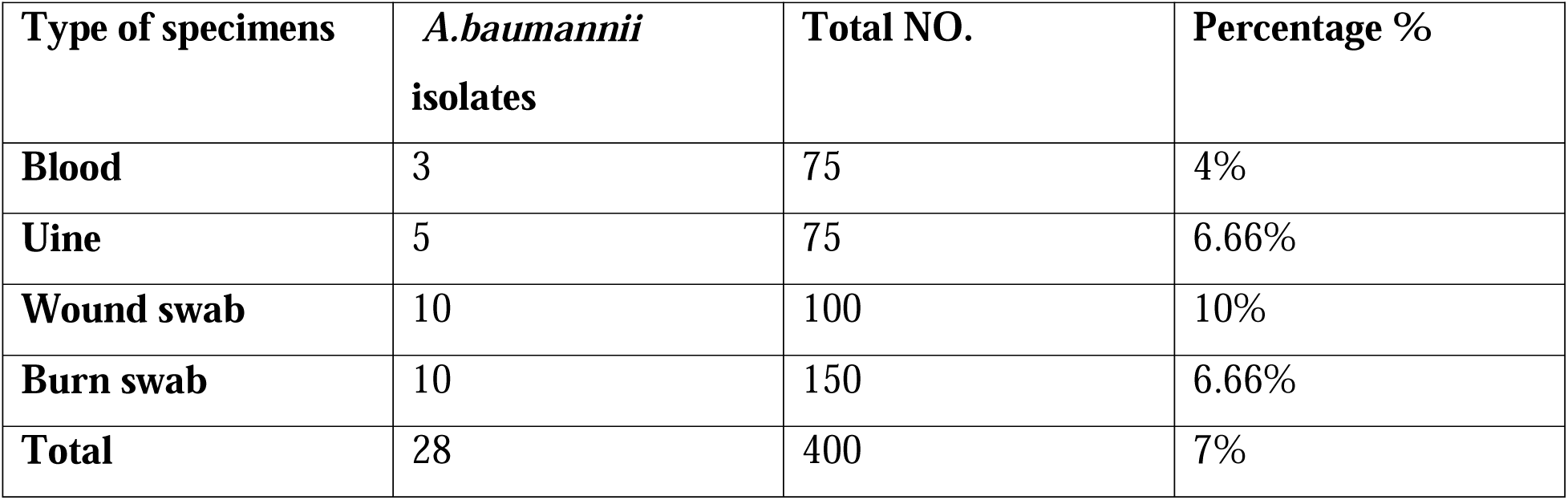
Percentage of specimens used in this study.

In the realm of Antibiotic Susceptibility Testing (AST) for *Acinetobacter baumannii,* an examination of 28 isolates unveiled diverse patterns of antibiotic resistance and sensitivity, as depicted in Figure (1). The findings revealed that 81.2% and 80% of the isolates displayed resistance to carbenicillin and piperacillin, respectively. Concerning β-lactam/β-lactamas inhibitor combination agents, 60% of the isolates exhibited resistance to piperacillin-tazobactam. Resistance rates to third-generation cephalosporins were documented as follows: cefotaxime (70.2%), ceftriaxone (71.6%), and ceftazidime (70.2%). Additionally, 65.8% of the isolates demonstrated resistance to the fourth-generation cephalosporin, cefepime. These results underscore the diverse antibiotic resistance profiles manifested by the *A. baumannii* isolates.

**Figures (1):**
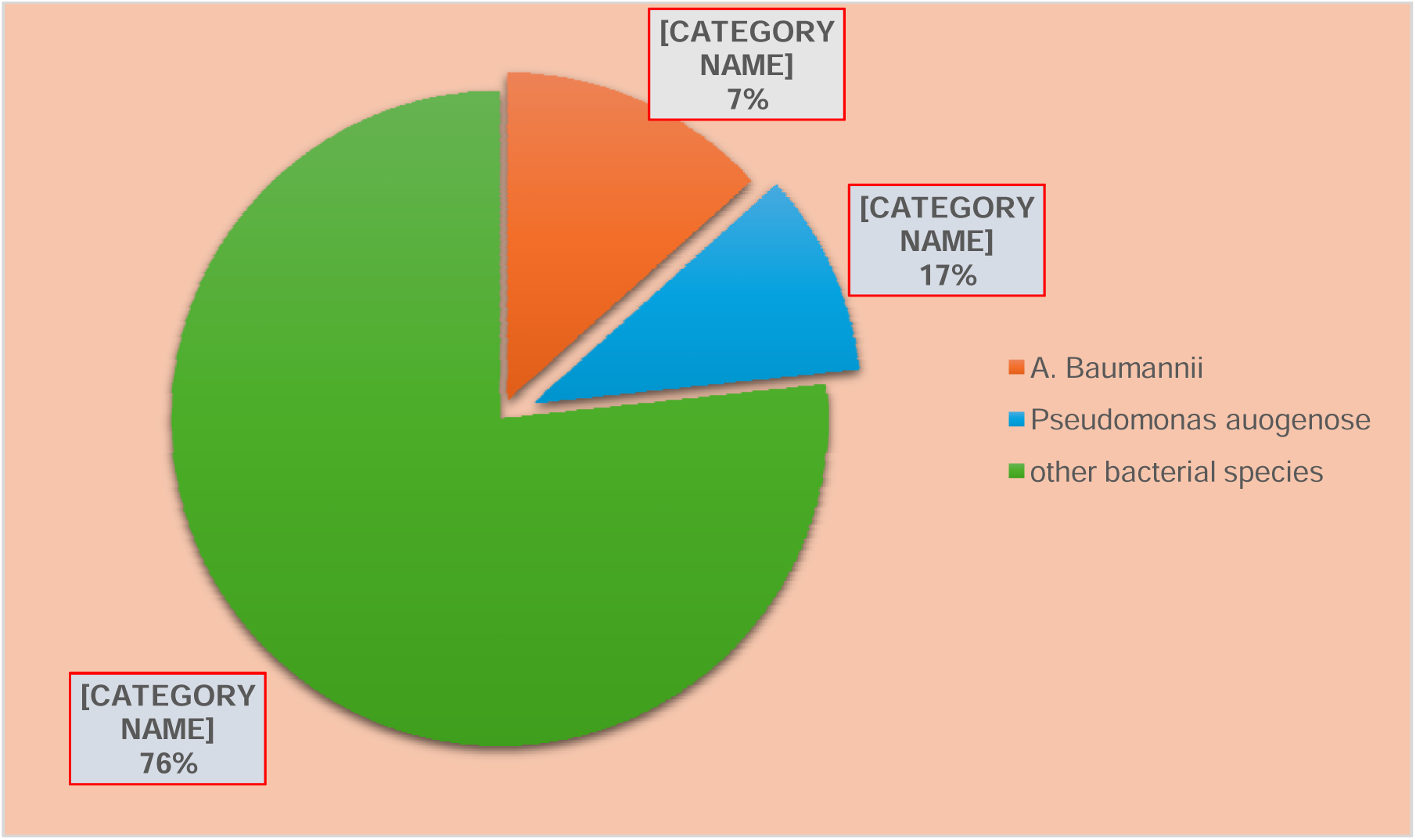
Distribution Isolation and identification of *A.baumannii* from Different Clinical specimens.

**Figure (2):**
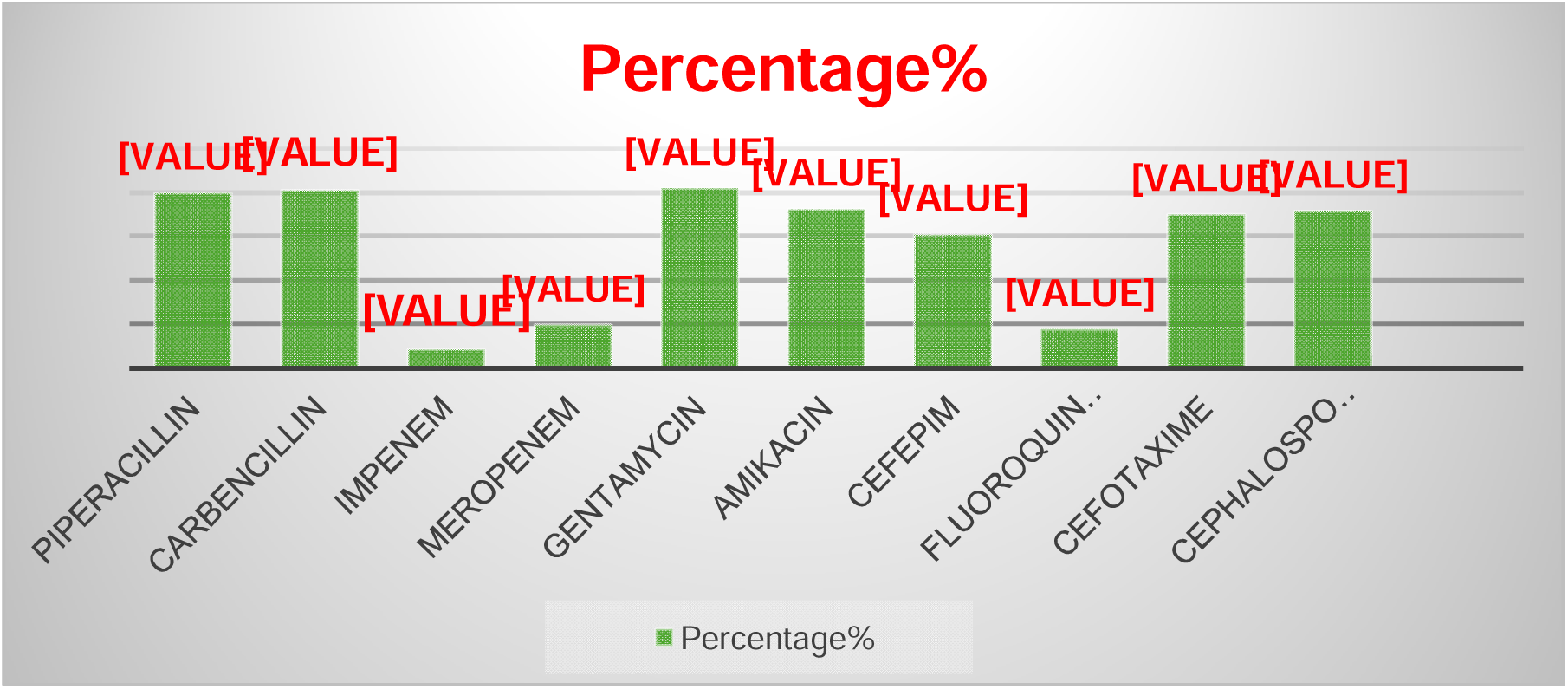
Antibiotic Susceptibility Test of *A.baumannii* to isolates toward Antibiotics (No. 28).

Resistance to Imipenem and meropenem among the isolates was noted at rates of 8.4% and 19.8%, respectively. Aminoglycoside antibiotics exhibited an overall resistance percentage, with the highest levels observed in gentamicin (82%) and amikacin (72.2%). Regarding fluoroquinolones, 80.4% of the isolates demonstrated resistance to ciprofloxacin. Antibiotic resistance was identified across various classes, including multi-drug resistance (MDR), extensive drug resistance (XDR), pan-drug resistance (PDR), and resistance to aminoglycosides, carbapenems, cephalosporins, β-lactam/β-lactamase inhibitor combinations, quinolones, monobactam, and antibacterial penicillin. The *A. baumannii* isolates’ susceptibility to antibiotics was evaluated using the Kirby-Bauer disk diffusion method. 28 (7%) of the isolates tested positive for multiple drug resistance tests. 80% of the isolates showed piperacillin resistance during the investigation. Antibiotic resistance seems to have increased based on earlier reports from Al-Hilla Teaching Hospital [6]. The piperacillin susceptibility of *A. baumannii* can be reported in a few different ways, with reported values ranging from 31.7% to 64.5. Similarly, research, also called carboxypenicillins—show higher variability. It is effective against ampicillin-resistant *A. baumannii* conducted through study by [7], the isolated *A. baumannii* strains showed a moderate rate of resistance to piperacillin. The class of carbenicillins In an new study, Tests in Al-Hilla showed that carbenicillin performed better [8].7% of the isolates in the current study showed drug resistance, despite piperacillin /tazobactam being found to be moderately active against *A. baumannii* isolates. The best antimicrobial agent against *A. baumannii*, according to several studies, is piperacillin-tazobactam [9].This study’s cefepime resistance rate of 60.8% was greater than the 50% reported in a different local study [10]. Fourth-generation antibiotic cefepime exhibits enhanced resistance against β-lactamases and an extended spectrum, especially AmpC β-lactamases, and a heightened capacity to permeate the porins found in the outer membrane of Gram-negative bacteria. The medication carbapenems is expensive and not easily accessible in the province of Hilla.. However, Numerous recent studies have reported the high activity of this class of antibiotics against *A. baumannii.* Furthermore,imipenem was found to be very active against *A. baumannii* in a number of past studies [10]. The activity of carbapenems, specifically meropenem and imipenem, against *A. baumannii* is the subject of this study. According to the results, which are consistent with earlier research, meropenem is more active than imipenem, with resistance rates of 22.4% and 20.4%, which are in disagreement with [12] and agreement with that published by [13], who found that meropenem was better than imipenem. The isolates of *A. baumannii* found in the hospitals in Hilla appear to be less susceptible to fluoroquinolones due to increased or cumulative use of the medication. Susceptibility to fluoroquinolones appears to be decreasing faster (17.6%) than susceptibility to other antimicrobial classes, according to a comparison with earlier studies done in Hilla. Multiple antibiotic resistance was present in every *A.baumannii* isolate used in this investigation. Still, as was to be expected, disparities in MDR definitions precluded any meaningful comparison of the variations in MDR rates among studies other than the current one. *A. baumannii* Carbapenem-Resistant The initial screening test for carbapenem susceptibility was applied to all 28 isolates of *A. baumannii,* adhering to the guidelines for initial screening for carbapenem susceptibility provided by the Clinical and Laboratory Standards Institute. using the disk diffusion method and antibiotic disks containing 10μg of meropenem and imipenem each. According to CLSI,2023 recommendations, they were classified as resistance for Imipenem and meropenem if the inhibition zone diameter was ≥19 mm, intermediate if it was 16–18 mm, and resistant if it was ≤15 mm. Figure (2) lists the isolates’ susceptibilities to imipenem and meropenem based on the results of the susceptibility tests. Throughout the study period, Meropenem had. The activity (19.8%) higher than Imipenem (8.4%). However, Among the 28 isolatesdisplaying resistance to both imipenem and meropenem, 23 exhibited cross-resistance to both antibiotics. The study also investigated the carbapenemase-producing capability of these *A. baumannii* isolates, which were already resistant to carbapenem. Employing both phenotypic and molecular techniques, including Polymerase Chain Reaction (PCR), the researchers assessed the potential of *A*. *baumannii* isolates falling into classes A (serine-βlactamases) and B (metallo-βlactamases, MBL) to produce carbapenemase. The PCR method played a crucial role in identifying carbapenemase production in *A. baumannii.* The study unveiled that *A. baumannii’s* resistance to carbapenemase is often influenced by various factors. These include the loss of proteins in the outer membrane, synthesis of Metallo-β-lactamases (MBL) and carbapenemase, or the regulation of active efflux pumps. Notably, MBLs have become more prevalent in clinical isolates globally in recent years, and strains producing these enzymes have been linked to severe infections and prolonged nosocomial outbreaks(14). The tests using PCR could be used in Hilla hospitals to determine whether decreased susceptibility to carbapenemase is associated with the production of carbapenemases (15). By using PCR amplification to identify isolates with carbapenemase genes, this study explored the phenotypic and molecular genes of carbapenemase. The study specifically concentrated on genes linked to metallo-beta-lactamases (MBLs), such as *bla_VIM_*and *bla_KPC_*. Further information about the kinds of MBLs found in the 40 isolates that tested positive for *A. baumannii* was obtained using the Kirby-Bauer disk diffusion technique.The findings showed that all isolates (100%, 28/400) had *bla_-OXA-51_* genes, with *bla_VIM_*being the most common in 12 cases (42.8%) and *bla_KPC_* in 10 cases (35.7%). The difficulty in combating resistance mechanisms is highlighted by their high prevalence, especially *bla_VIM_*-type genes. highlights the difficulty of using beta-lactam antibiotics to treat *A. baumannii* infections. For the purpose of creating efficient treatment plans and reducing the effects of antibiotic resistance, it is essential to comprehend and keep an eye on patterns of antibiotic resistance, particularly the frequency of particular MBL genes.the spread of strains resistant to multiple drugs (16).

**Figure (6):**
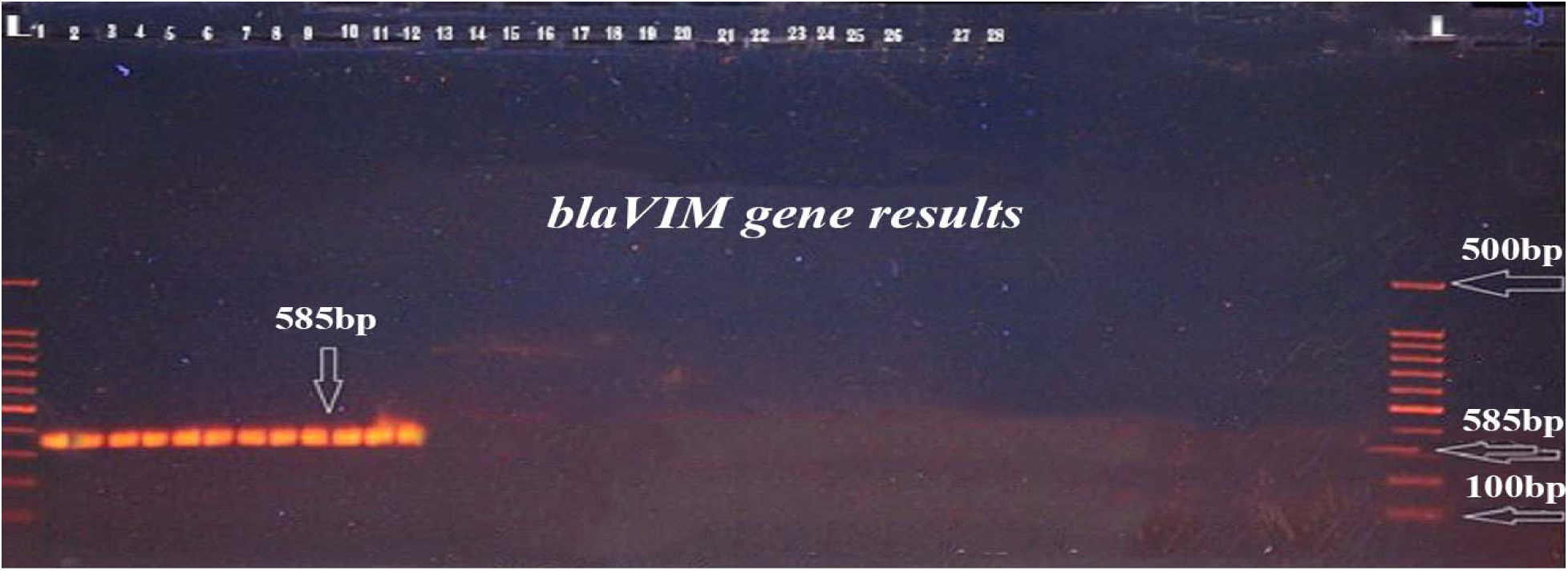
The electrophoreses of PCR amplified products of *bla_VIM_* gene was performed at 60 volts for 2 hr. (100-500bp ladder). Lanes (1-12) of *A.baumannii* isolates show positive results with *bla_VIM_*(585bp)

**Figure 8:**
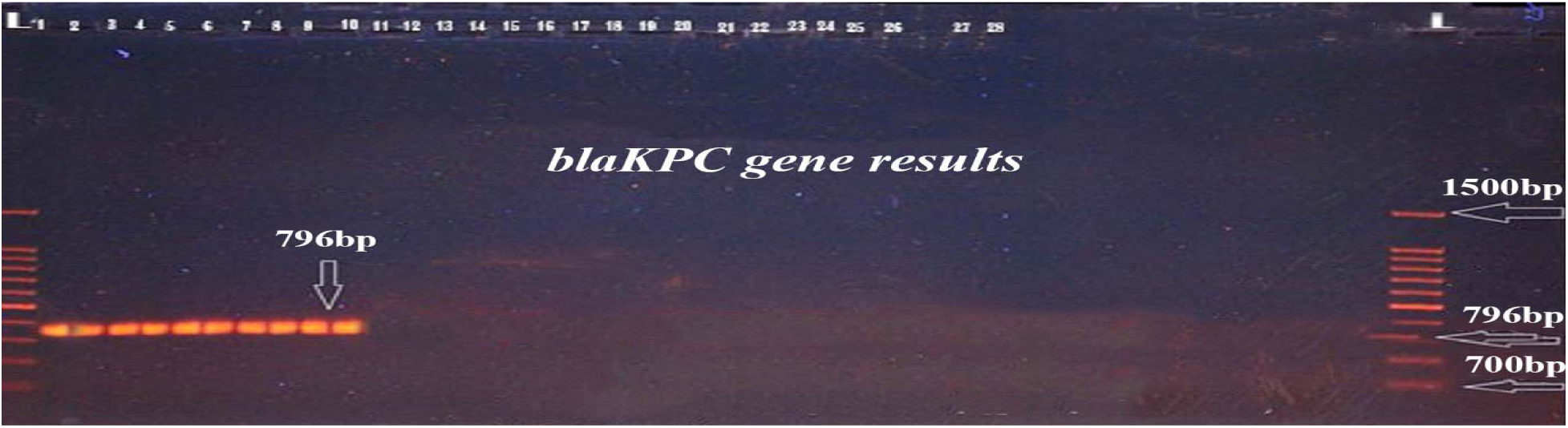
The electrophorese of PCR amplified products of *bla_KPC_* gene was performed at 60 volts for 2 hr. (700-l500bp ladder), Lanes (1-10) of *A.baumannii* isolates show positive results with *bla_KPC_* (796 bp).

**Figure 9:**
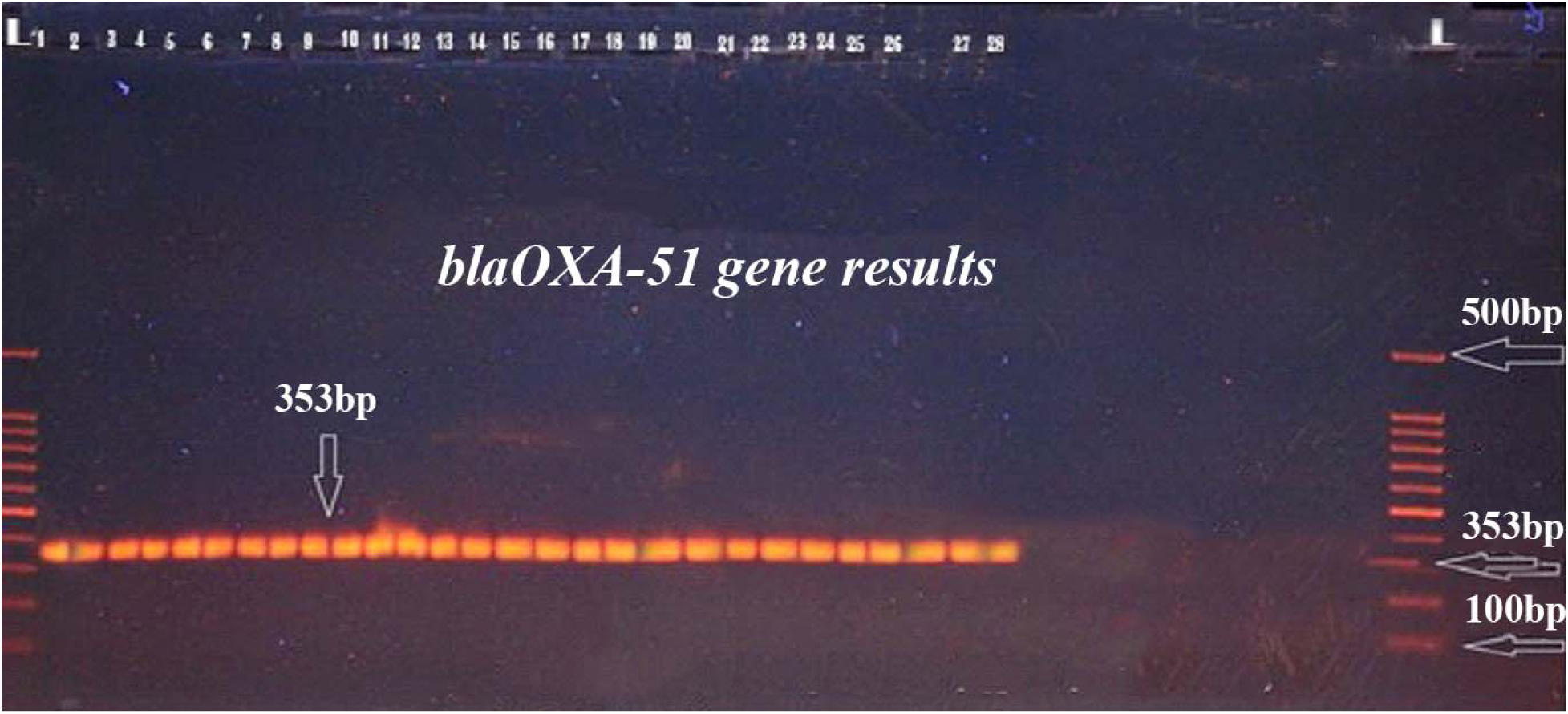
The electrophorese of PCR amplified products of *bla_OXA-51_gene* was performed at 60 volts for 2 hr. (100-500bp ladder), Lanes (1-28) of *A.baumannii* isolates show positive results with *bla_OXA-51_gene* (353 bp).

**Table 3:**
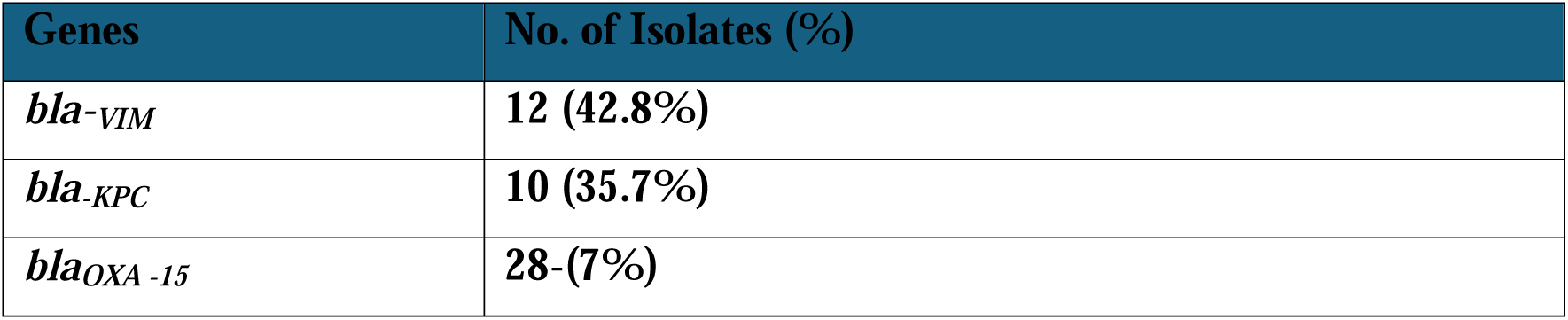
Distribution of Carbapenemase genes in 28*A.baumannii* isolates.

The production of Metallo-beta-lactamases (MBLs) by *Acinetobacter baumannii* presents a serious epidemiological risk since these enzymes can confer resistance to a wide range of beta-lactam antibiotics (17). These antibiotics are hydrolyzed by MBLs, which limits the range of possible treatments and makes the antibiotics ineffective. The multidrug-resistant phenotype of *A. baumannii* is a result of its acquisition and expression of MBLs, which makes treating infections brought on by this opportunistic pathogen more difficult.The risk associated with epidemiology stems from multiple factors (18). First of all, *A. baumannii* is well-known for its adaptability and capacity to endure in medical environments, which can result in nosocomial infections. It becomes more difficult to contain outbreaks in healthcare facilities as a result of MBL development, which expands its repertoire of resistance. Second, the possibility of horizontal gene transfer makes it possible for MBL genes to spread quickly among various bacterial strains, aiding in the spread of resistance to antibiotic. The emergence of MBLs in *A. baumannii* underscores the urgency of robust surveillance, infection control measures, and antibiotic stewardship to curb the dissemination of multidrug-resistant strains(20). Addressing this epidemiological risk requires a multifaceted approach that includes prudent antibiotic use, enhanced hygiene practices, and the development of novel therapeutic strategies to combat drug-resistant *A. baumannii* infections. 9 –MBL have been known in Gram-negative bacilli [21]., In *A. baumannii,* carbapenemase and metallo-beta-lactamases (MBLs) are commonly identified as KPC and VIM types. A technique called PCR amplification was used to find these MBL genes. Polymerase Chain Reaction (PCR), a molecular technique, made it possible to specifically identify isolates that carried MBL genes, such as *blaVIM* and *blaKPC* for carbapenemase. The identification process’s use of PCR represents an accurate and effective way to determine whether these resistance-conferring genes are present in *A. baumannii* isolates, offering vital data for comprehending the frequency and distribution of MBLs and informing management plans for antibiotic resistance in healthcare settings across the globe (22).

28/400(7%) of the 28 *A. baumannii* isolates that were carbapenem-resistant had the oxallinase genotype. The finding implies that *blaVIM* was the most prevalent carbapenemase-type gene in national hospitals. Similar results were also observed in Japan and Italy, *A. baumannii* strains that produce VIM β-lactamase proliferated as hospital pathogens [23]. There are currently 23 members of the VIM family [24], occurring mainly in *A.baumannii* within multiple-integron cassette structures. Of the 28carbapenemresistant isolates, 28 (7%) had *blaOXA-51* genes verified by PCR analysis. The results of this investigation indicate that, among the isolates of *A. baumannii* studied in this study, *blaKPC* is indeed a gene that encodes Carbapemase, specifically in the context of antibiotic resistance in *Acinetobacter baumannii*. This gene, along with others such as *blaVIM* and *blaKPC,* contributes to the ability of bacteria to hydrolyze and inactivate a broad range of beta-lactam antibiotics, posing challenges for effective treatment.When investigating the presence of MBLs, particularly in *A. baumannii*, Detecting the *blaKPC* gene is pivotal, and leveraging PCR amplification in the identification process offers precision and specificity. This targeted approach not only aids in discerning antibiotic resistance patterns but also provides valuable insights for implementing effective infection control measures and refining treatment strategies within healthcare settings (26).

## Conclusion

The combined genotypic and phenotypic detection of carbapenemases in clinical isolates of *A. baumannii* is essential for a holistic approach to antibiotic resistance management. Identifying key resistance genes in Acinetobacter baumannii, such as *bla_OXA-51_* while carbapenemase including *bla_VIM_*, and *bla_KPC_*-type have the ability for hydrolysis of B-lactam antibiotic.This knowledge contributes to the development of targeted treatment strategies, facilitates infection control measures, and ultimately helps preserve the effectiveness of antibiotics in the face of emerging resistance. Continued research in this area is vital to stay ahead of evolving resistance patterns and to inform public health efforts.

## Data Availability

CLINICAL SPECIMENS

